# “It hurts your heart”: frontline healthcare worker experiences of moral injury during the COVID-19 pandemic

**DOI:** 10.1101/2022.06.17.22276433

**Authors:** Siobhan Hegarty, Danni Lamb, Sharon A. M. Stevelink, Rupa Bhundia, Rosalind Raine, Mary Jane Doherty, Hannah R. Scott, Anne Marie, Victoria Williamson, Sarah Dorrington, Matthew Hotopf, Reza Razavi, Neil Greenberg, Simon Wessely

## Abstract

**Background:** Moral injury is defined as the strong emotional and cognitive reactions following events which clash with someone’s moral code, values or expectations. During the COVID-19 pandemic, increased exposure to potentially morally injurious events (PMIEs) has placed healthcare workers (HCWs) at risk of moral injury. Yet little is known about the lived experience of cumulative PMIE exposure and how NHS staff respond to this.

**Objective:** We sought to rectify this knowledge gap by qualitatively exploring the lived experiences and perspectives of clinical frontline NHS staff who responded to COVID-19.

**Methods:** We recruited a diverse sample of 30 clinical frontline HCWs from the NHS CHECK study cohort, for single time point qualitative interviews. All participants endorsed at least one item on the 9-item Moral Injury Events Scale (MIES) (Nash et al., 2013) at six month follow up. Interviews followed a semi-structured guide and were analysed using reflexive thematic analysis.

**Results:** HCWs described being routinely exposed to ethical conflicts, created by exacerbations of pre-existing systemic issues including inadequate staffing and resourcing. We found that HCWs experienced a range of mental health symptoms primarily related to perceptions of institutional betrayal as well as feeling unable to fulfil their duty of care towards patients.

**Conclusion:** These results suggest that a multi-facetted organisational strategy is warranted to prepare for PMIE exposure, promote opportunities for resolution of symptoms associated with moral injury and prevent organisational disengagement.

**Highlights:**

- Clinical frontline healthcare workers (HCWs) have been exposed to an accumulation of potentially morally injurious events (PMIEs) throughout the COVID-19 pandemic, including feeling betrayed by both government and NHS leaders as well as feeling unable to provide duty of care to patients
- HCWs described the significant adverse impact of this exposure on their mental health, including increased anxiety and depression symptoms and sleep disturbance
- Most HCWs interviewed believed that organisational change within the NHS was necessary to prevent excess PMIE exposure and promote resolution of moral distress

## Introduction

The global COVID-19 pandemic continues to expose healthcare workers (HCWs) to an increased degree of occupational stress. Since March 2020, high levels of depression, anxiety disorders, Post-traumatic stress disorder (PTSD) and sleep disorders have been reported among healthcare workers in the U.S, China and Europe (e.g. Greene et al., 2021; Lamb et al., 2021; Marvaldi et al., 2021; Roberts et al., 2021). HCWs also face a high risk of exposure to potentially morally injurious events (PMIEs), which include ‘perpetrating, failing to prevent, bearing witness to, or learning about acts that transgress deeply held moral beliefs and expectations’ (Litz et al., p.700). Exposure to PMIEs has been identified as a trigger for what has been termed moral dissonance (Farnsworth et al., 2017). This occurs when an event violates a person’s beliefs about right and wrong, the person will experience internal dissonance between beliefs, i.e. “I am a good person” but “I did something unforgivable”. Left unresolved, this value compromise can manifest as moral injury, involving deep feelings of guilt, shame, and a sense of betrayal (Barnes et al., 2019), anger and moral disorientation (Molendijk, 2018). This in turn has been associated with adverse psychiatric, social, functional, and spiritual consequences (Farnsworth et al., 2017). Another related construct that has been discussed in relation to HCWs is that of moral distress, which is typically a short-term, reversible reaction to PMIEs; this occurs when providers believe they are doing something ethically wrong and have little power to change the situation (Jameton, 1993). In this instance, the imposed action indicates the moral distress is caused by factors external to HCWs (Bruce, 2015; Wiegand, 2012).

Whilst moral injury does not constitute a diagnosable mental disorder (APA, 2020), a systematic review and meta-analysis has found that it is significantly associated with PTSD, depression, and suicidal ideation across a range of professions (veterinarians, teachers, police officers, journalists, military personnel) and countries (Williamson et al 2018).

By nature of their occupation, many HCWs are routinely exposed to morally challenging situations, including having to make decisions which may lead to patients dying or failing to recover from serious conditions. Even prior to COVID-19, clinician burnout was an increasing concern (Reith, 2018), with evidence suggesting nurses in England have elevated rates of burnout compared to peers from other parts of Europe (Aiken et al., 2014). During the pandemic, reports of healthcare providers facing increased pressure to see more patients (Sibeoni et al., 2019) have been common, despite understaffing and insufficient resources (Greenberg et al., 2020). Unsurprisingly, experts in the field have suggested these experiences may constitute PMIEs among HCWs (Best, 2021).

Estimates of the prevalence of moral injury in HCWs responding to the pandemic range from to 32% in the US (Rushton et al., 2021), to between 20 and 41% in China (Zhizhong et al., 2020: Wang et al., 2021). One global study found that 27% of HCWs experienced moderate impairment in social or professional functioning related to moral injury symptoms, which increased to 46% at phase two of the study, conducted six months later in a separate, larger sample (Mantri et al., 2021). Furthermore, Wang and colleagues (2021) found that moral injury symptoms were strongly associated with higher clinician burnout, greater psychological distress, and lower level of subjective well-being.

Qualitative work conducted early in the pandemic (March-May 2020) highlighted several potential indicators of resilience to moral injury: honest and timely communication delivered by trusted leaders, sufficient Personal Protective Equipment (PPE) and visible efforts by management to protect staff, proactive effort to remain connected to those working from home, opportunity for rest and recovery and preparing staff for new tasks, collaborative decision making, for instance by jointly finding solutions to ethical problems (Kreh et al., 2021). However, the nature and impact of cumulative PMIE experiences during the ongoing COVID-19 pandemic remain unclear; little work has been done to explore how HCWs respond to PMIEs, nor how organisations can support staff in preventing and mitigating the effects of PMIEs. Given that PMIE experience is subjective to the individual, and the lack of validated measure to assess moral injury, the current study adopted a qualitative approach to explore PMIEs.

We explored the experiences and impact of PMIEs on HCWs wellbeing, as well as HCWs beliefs about organisational practices that influence exposure to, and wellbeing outcomes following PMIEs.

## Methods

### Design and overview

We nested a single time point qualitative interview study within NHS CHECK, a prospective cohort study examining the health and wellbeing of HCWs during the COVID-19 pandemic, full details of which are outlined in our protocol paper (Lamb, Greenberg, et al., 2021). We purposively sampled 30 clinical HCWs who self-reported PMIE exposure in the NHS CHECK six month follow up survey, for inclusion in this study. The interviews were analysed using reflexive thematic analysis, informed by a critical realist approach to inquiry (Bhaskar, 1975, 1977, 1994). Ethical approval for the study was granted by the Health Research Authority (reference: 20/HRA/2107, IRAS: 282686) and local Trust Research and Development approval.

### Participants

Participants were NHS affiliated staff, drawn from a pool of NHS CHECK respondents who consented to be contacted again by the study team. NHS CHECK is an MRC-funded cohort study, involving 18 (acute and mental health) NHS Trusts across England. All staff in participating Trusts were emailed an invitation to complete the on-line survey. To date we have collected quantitative survey data from 23,462 participants at three time points from (baseline, 6-month and 12 month follow up). The full NHS CHECK cohort is broadly representative of the wider NHS workforce demographics (NHS Digital, 2021). Participants from the NHS CHECK 6-month cohort who consented to be recontacted were purposively sampled for this qualitative study. All participants endorsed at least one item on the 9-item Moral Injury Events Scale (MIES) (Nash et al., 2013) (moderately agree/agree/strongly agree) in the NHS CHECK six month follow up survey. For context, the prevalence of self-reported PMIE exposure in the overall NHS CHECK sample was 14.6% at baseline, and 26.8% at six months (Lamb et al., Under review February 2022, OEM). Sampling was conducted to ensure diversity across both exposure to moral injury sub-types (i.e. moral injury by commission, by omission, by betrayal, and some mix of these), and a mix of those reporting adverse mental health symptoms and those not reporting such symptoms. Variation was sought in mental health outcomes and PMIE exposure type to be as inclusive as possible of the wide range of HCW experiences. Additionally, the sample is highly demographically diverse (NHS trust, age, sex, ethnicity, clinical occupation).

We used the Patient Health Questionnaire (PHQ-9) (Kroenke et al., 2001) to indicate probable depression, which is scored on a scale of (0-27; where 0-4 indicates none to minimal, 5-9 mild, 10-14 moderate, 15-19 moderately severe, 20-27 severe). The General Anxiety Disorder (GAD-7) (Spitzer et al., 2006) was used to indicate probable anxiety, scored on a scale of (0-21; where 0-4 indicates minimal, 5-9 mild, 10-14 moderate, 15-21 severe). The civilian version of the PTSD Checklist (PCL-6) (Lang & Stein, 2005) was used to indicate probable PTSD, scored on a scale of (6-30; with a score of 14 of above indicating probable PTSD). We recruited fourteen participants whose scores on the above measures were consistent with little to no adverse mental health symptoms (i.e. scoring 0-4 on PHQ-9, 0-4 on GAD-7, 6-9 on PCL-6). We recruited 16 participants whose scores met clinical threshold for at least one of the above probable disorders (i.e. scoring ≥ 10 on PHQ-9, and/or ≥10 on GAD-7, and/or ≥ 14 on PCL-6).

### Procedure and materials

Eligible participants were contacted via the email addresses they provided in the NHS CHECK survey. The initial recruitment email invited potential participants to share their experiences in a qualitative interview, and included the participant information sheet (PIS), which clarified that participation in the study was voluntary and that no identifiable data would be made available outside of the immediate research team. In total, 239 HCWs-from 14 NHS Trusts spread geographically across England-were sent an invitation to interview. 30 participants were selected from a greater number who expressed interest to maximise diversity of the sample. Staff from 12 of the 14 trusts invited responded to the initial outreach email. The selected sample (comprising staff from these 12 trusts) were sent a follow up email including a Calendly link to book a time and date for interview with a researcher. Two days prior to interview, participants were sent a reminder email including a link to a digital consent form. The interview followed a semi-structured topic guide (see Appendix A) and focused on the following key areas: the nature of PMIEs experienced, their impact on mental health, how staff have dealt with PMIEs including personal and work-based sources of support they found helpful/unhelpful to manage them, as well as what Trusts can do to prevent/mitigate PMIEs. All 30 HCWs sampled completed the interview in full and were compensated for their time with a £25 gift voucher.

Interviews took place between the 17^th^ November 2021 and 14^th^ of December 2021 and lasted between 27 minutes and 1 hour and 3 minutes (mean 43 minutes). Interviews were professionally transcribed verbatim, with identifying information removed. Pseudonyms are used throughout this paper.

### Data Analysis

We followed the principles of reflexive thematic analysis (Braun & Clarke, 2019, 2021) throughout this study. The first author was immersed in the data by reading and rereading all the transcripts, reflecting on the interviews, and discussing the development of themes with the wider research team. A critical realist approach was adopted to understand the perspectives and experiences of participants throughout. All coding was inductive, derived from the data and not determined by pre-existing theories. The first five transcripts were provisionally coded and uploaded along with the remaining transcripts to NVivo 12 for Windows. Subsequent transcripts were coded into this framework, which was revised and expanded throughout the coding process. Similar codes were combined to explore commonalities between lower-level codes. The final stage of analysis involved conceptual linking of themes which was achieved through discussion with the wider research team, and through consideration of feedback on drafts of the evolving analysis.

### Reflexivity

The research team behind this study comprised a diverse group of researchers, including different genders, career stages, and clinical specialties. The first author (S.H.) is an early career researcher and recent MSc graduate of Clinical Mental Health Sciences. S.H. was deeply immersed in the data and analysed it with regular input from the wider research team. S.H. kept a reflexive journal to highlight the feelings and thoughts aroused by the analytic process and bring awareness to how they shaped theme generation. S.H. debriefed her thought process surrounding coding to D.L., who is a senior research fellow with over a decade of experience of mental health research and is a co-investigator on NHS CHECK. In the early stages of analysis, D.L. checked various stages of coding and theme development. To enhance trustworthiness and credibility S.H. discussed theme development with the wider research team (consisting of clinically active NHS staff and non-clinical researchers) to bring awareness to blind spots in data interpretation. Through reflective discussion S.H. became aware of the systemic lens through which she had initially interpreted participants’ experiences. This prompted S.H. to revisit the transcripts with a view to understand how PMIEs were processed on an individual level. This allowed insight into individual differences in cognitive appraisals captured, providing a more holistic view of the data. The expertise of the research team enabled a deep insight into this subject. However, there are potential disadvantages in our closeness to the topic. We sought to address this throughout data analysis by maintaining curiosity about our data, discussing and welcoming alternative views to interpretation, and regularly comparing the evolving analysis to the raw data.

## Results

### Descriptive information

30 HCWs volunteered and took part in the study, out of 239 approached. The socio-demographic and occupational characteristics of the participants are shown in Table 1, as well as their PMIE exposure type and mental health status. All participants had experienced changes in their work practice and several were redeployed. Almost all participants had face to face contact with colleagues and patients in hospital settings, though some participants (typically mental health professionals) worked in the community or remotely from home.

**Table 1.** Demographic characteristics of sample (n =30)

| Characteristic | N(%) |
| --- | --- |
| Gender |  |
| Male | 10 (33.3%) |
| Female | 20 (66.7%) |
| Occupation |  |
| Occupational Therapist | 1 (3.3%) |
| Speech and Language Therapist | 1 (3.3%) |
| Psychological Therapist | 3 (10%) |
| Radiographer | 1 (3.3%) |
| Physiotherapist | 1 (3.3%) |
| Nurse | 15 (50%) |
| Doctor | 2 (6.7%) |
| Pharmacist/Pharmacy Technician | 3 (10%) |
| Midwife | 1 (3.3%) |
| Healthcare Assistant | 2 (6.7%) |
| Ethnicity |  |
| White | 18 (60%) |
| Asian | 6 (20%) |
| Mixed | 3 (10%) |
| Black | 2 (6.7%) |
| Other | 1 (3.3%) |
| Mental Health Status * |  |
| GAD-7 case ($\geq 10$) | 10 (33.3%) |
| PHQ case ($\geq 10$) | 14 (46.7%) |
| PCL case ($\geq 14$) | 14 (46.7%) |
| No to mild symptoms <sup>a</sup> | 14 (46.7%) |
| PMIE exposure type |  |
| Omission | 16 (53.3%) |
| Commission | 9 (30%) |
| Betrayal | 24 (80%) |
| * Can meet threshold for >1 probable mental health condition. In total 53.34% of participants met criteria for any mental disorder. <sup>a</sup> No to mild symptoms indicates scoring 0-4 on PHQ-9, 0-4 on GAD-7, and 6-9 on PCL-6. |  |

### Finding Summary

The majority of participants described either feeling ill equipped and/or under-supported to respond to crisis. Participants commonly linked chronic, pre-pandemic, government underfunding of the NHS to several systemic organisational issues such as understaffing, which, in turn, strained the working relationships between staff and management. Exacerbations of these issues during the pandemic (e.g. teams that were already short-staffed lost additional members to sick leave and isolation) led to feelings of betrayal, as staff felt limited in the support they could offer patients (e.g. where no pathway was available for referral) and sometimes felt forced to act outside their clinical competencies. This left participants feeling unable to fulfil their duty of care to patients. In response to PMIEs, staff could sometimes avoid the experience of moral dissonance. However, in many situations this was not possible and instead, the experience of moral distress took a considerable psychological toll, leading some to consider leaving their role or trust. Staff sought to adaptively manage their moral distress, and occasionally were successful in resolving it; yet, for most this was depicted as an on-going struggle that would require organisational change to tackle. These issues are summarised in the following table.

**Table 2.**
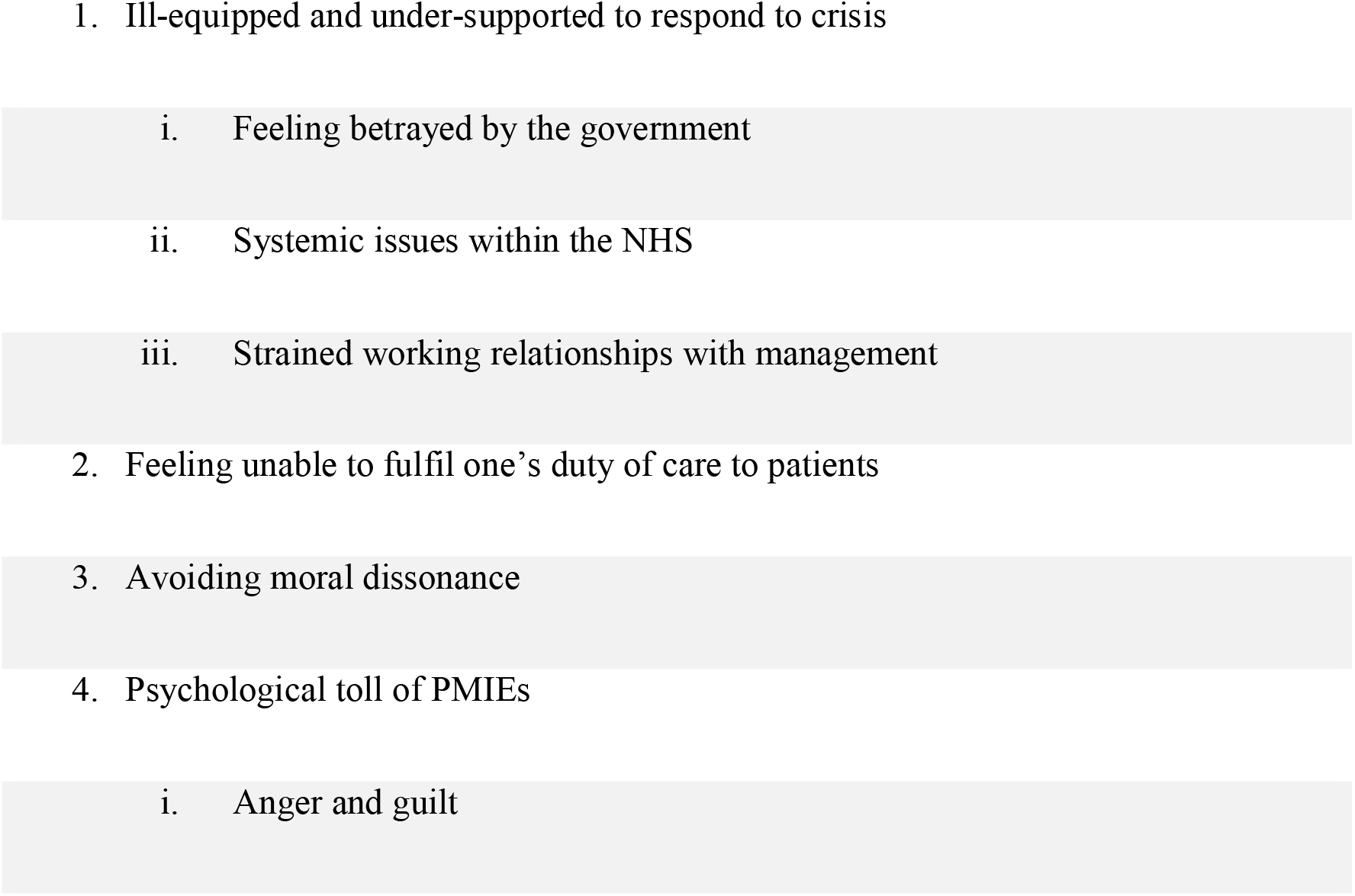
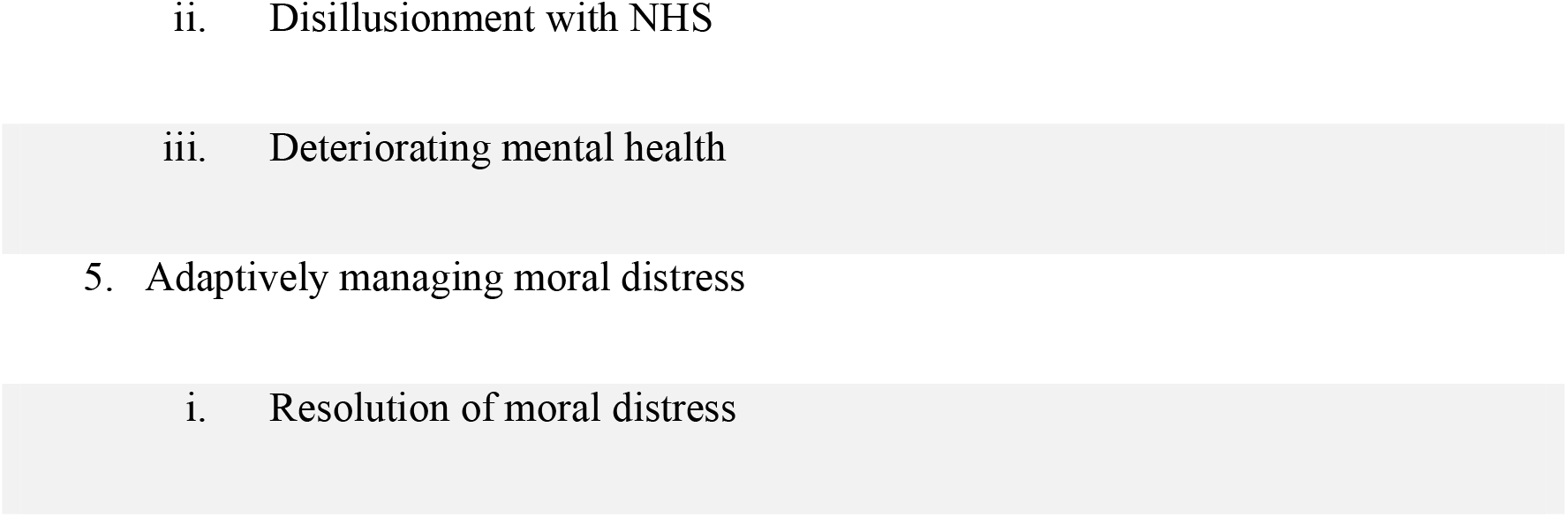
Themes and sub-themes

## 1. Ill-equipped and under-supported to respond to crisis

### 1.i. Feeling betrayed by the government

Almost half of participants expressed feeling betrayed by the government due to perceived mismanagement of national regulations surrounding COVID-19, compounding existing disappointment with perceived chronic government underfunding of the NHS. Staff were angered by regular changes of guidelines, which often were not felt to reflect the severity of the situation nor perceived to be based on scientific evidence. Participants had differing views on the severity of regulations.

Most participants who felt betrayed by the government, felt rules were inappropriately lax during time periods with the highest number of COVID-cases in hospitals.

> *The fact that they [the Government] effectively mock the work that we’re doing by saying that you’ve now got your freedom back and it’s basically saying that there isn’t a virus that’s spreading.* (Harry, Nurse)

While a small minority felt the duration of lock downs were disproportionate to COVID threat.

> *I just think the way it was handled by the Government was so damaging overall and that’s going beyond the National Health Service. That’s including people’s mental health, suicide levels, the economic cost on people, the increase in domestic abuse.* (Melanie, Nurse)

These views deepened pre-existing distrust in the government’s willingness to protect the interests of the NHS and its staff.

### 1.ii. Systemic issues within the NHS

This sub-theme encapsulates pre-existing systemic issues, felt to contribute to staff feeling ill equipped and under-supported by the NHS during the pandemic.

A perceived lack of funding to the NHS was felt by participants to increase their exposure to situations of moral conflict. HCWs on wards routinely found themselves understaffed on shifts, with inadequate medical equipment to serve the volume of patients. Additionally, several HCWs struggled with organisational regulation of scarce PPE, including inequitable distribution and delays mandating PPE use; both situations were viewed to put staff and non-COVID patients at risk.

> *It was all about, well if you are doing frontline care to someone with COVID then you can have PPE but absolutely no sense about trying to protect anyone else.* (Gwen, Midwife).

While PPE shortages were less frequent following the first wave, staffing and other resource shortages were described as the norm across settings.

> *What is the measure of a hospital being overwhelmed? Is it that there are no beds for the patients, is it that there are no staff to look after the patients because we’ve been in all of those situations. Is it when you have to decide whether a patient should go to ITU [Intensive Care Unit] or whether you think there’s a chance they’ll die so you say actually no we can’t afford them to go to ITU because we’ve been in that situation* (Harry, Nurse)

Additionally, a few HCWs felt unsupported by the reporting channels intended to facilitate disclosure of malpractice and mistreatment at work. In some trusts this was perceived as a systemic failure to respond to staff feedback about issues affecting personal or patient welfare and initiate subsequent organisational change. This contributed to the impression-shared by many-that the organisation was not looking out for individual staff members.

### 1.iii. Strained working relationships with management

This theme explores the factors felt to strain HCWs relationships to management, including absence from the frontline, lack of honesty and transparency, and deprioritising staff and patient needs.

Most staff felt those in upper management of their trusts were detached from the realities of work on the frontline.

> *We’re putting our health on the line here and we’re trying to keep ourselves and our families safe and we’re not getting the feedback and information because all our managers didn’t come in.* (Shaunagh, Radiographer)

This was construed as an uncaring approach by an upper-management which communicated access and discharge targets ( relating to the number of patients to access/be discharged from NHS services in a given period of time) from afar, which were not modified to account for staffing and equipment constraints. HCWs felt especially unsupported when the risk of a given clinical situation appeared to be intentionally obfuscated by upper-management. For instance, when trusts ran out of PPE for staff, or staff were not informed when patients were COVID positive: ‘*I felt quite let down by what seemed to be the lack of-an inability to communicate clearly and honestly*’ (Brianna, Nurse). HCWs across both physical and mental health settings believed their needs and the needs of their patients had been deprioritised by senior management during the pandemic to meet admission and discharge targets *“because all you are being told is hurry up and discharge people”* (Brianna, Nurse). This created the impression that trust priorities resided in the ‘*NHS business model’* and maintaining public optics of ‘*winning the war on COVID’*, with staff feeling ‘*disposable’* as a result. Staff resented feeling that the onus was put on them as individuals to compensate for systemic failings, exploiting their sense of moral duty towards patients.

Collectively, such experiences tainted the perception of organisational attempts to support staff, leaving some to feel like such attempts were disingenuous ‘*tick box’* exercises. Notably, HCWs generally recognised the pressure that middle management were under to bridge the ‘*gulf’* between upper management and frontline staff*. “I think they’re stuck and it’s not from them, we know it comes from above them so it’s corporate management like strategic level”* (Oliver, Nurse). HCWs experiences with individual line managers varied, with some feeling their line managers were either unsupportive or unapproachable and others feeling their managers did everything they could to support their wellbeing in the circumstances.

Those with line-management responsibilities in the sample, shed light on the difficulty of balancing their staff’s needs with the organisational demands they were tasked to impose upon them. This created routine moral conflict linked to redeploying staff members into challenging situations. These interviewees relayed feeling guilt for not having adequate time or emotional energy to address the concerns of individual staff members.

> *I’ve got a member of staff whose mum died on ITU, she’s terrified. I’ve got various people with back injuries. I’ve got quite a few people with mental health issues pre-pandemic which have been worsened, so stress, anxiety. […] my team are quite vulnerable, and I know them quite well.* (Stephen, Nurse)

Line managers often described subjugating their own needs to care for their staff, for instance by working far beyond their hours, and not taking annual leave. One manager expressed feeling similarly unsupported by those above him in the management hierarchy: *“there’s nobody asking after me”* (Harry, Nurse).

## 2. Feeling unable to fulfil one’s duty of care towards patients

This theme encompasses the ways in which HCWs felt they let their patients down throughout the pandemic. This includes instances in which staff felt that patients were directly harmed or put at risk by their actions, as well as instances where staff felt limited in their capacity to help patients. The latter was experienced due to a lack of/limited patient access to services in the pandemic, feeling inadequately trained and/or equipped to meet patients’ needs, and feeling too time pressured and understaffed to provide patient-centred care.

> *It’s just generally substandard care that we’re giving all of the time now and it’s become accepted, it’s become the norm.* (Conor, Healthcare Assistant)

Commonly, redeployed staff on wards described how they were required to act outside the remit of their clinical competencies which led to some participants feeling that they had endangered patients due to inadequate infection control procedures. Ward staff were sometimes unable to facilitate family communication with dying patients or support patients to die in a dignified manner due to the number of patients requiring attention. Conversely, when hospitals were less pressured, several participants described internal conflicts associated with perceived misuse of COVID emergency protocols, such as visitation restrictions. For instance, one staff member described feeling complicit in a ward manager’s decision to send the two daughters of a dying elderly patient home, despite being the only patient to occupy the room. This shared experience of letting patients down is conveyed by the participant below:

> *Those things that you are asked about when you are interviewed about why you want to be a nurse and everyone says it’s because I want to care for people. When you are unable to do that because of the just general resources so human resources and capacity then that’s really, really hurtful so it really hits you, it hurts your heart.* (Harry, Nurse)

Those working with non-COVID patients experienced a degree of moral distress related to having their services or teams cut during the pandemic, or feeling forced to provide sub-standard care via telehealth, and often having no operational pathway of referral for patients in urgent need.

> *I had families crying down the phone to me saying I need some help and I did feel like we were letting them down especially the families with children who were presenting as quite severely autistic who were still waiting for a diagnosis. Our hands were tied.* (Claire, Speech and Language Therapist)

Similarly, resource shortages led to mandates to shorten the duration of mental health interventions, leaving psychotherapists feeling that their interventions were not adequate to address the increasingly complex patient presentations. Time-limiting service provision in this way led to moral conflict for mental health professionals who predicted this would exacerbate the ‘*revolving door of patients’* leaving and re-entering the system, that predated the pandemic. Additionally, mental health crisis response was compromised. One HCW discussed routinely having to send people who were sectioned under the mental health act to remote parts of the country due to local bed shortages.

> *[…] then comes the question of what good is this going to do the patient in their recovery if they’ve been placed hundreds of miles away from friends and family.* (Owen, Nurse)

Several HCWs mentioned feeling like they had failed patients that were not seen during the pandemic. For instance, one HCW recounted how one of her patients had a surgery delayed due to the pandemic, which resulted in the patient’s condition deteriorating to the point of terminal illness.

> *It hadn’t spread anywhere else, and it would have been able to take the cancer out and in essence cure her. But by the time that she had her surgery which must have been four or five months later they scanned her before she had her surgery and it had spread. She was palliative.* (Chloe, Nurse)

Many HCWs shared the concern that they were part of services that were increasingly letting vulnerable people slip through the cracks.

## 3. Avoiding moral dissonance

The experiences discussed in both themes of ‘Ill-equipped and under-supported’ and ‘Feeling unable to fulfil one’s duty of care to patients’ capture a wide range of PMIEs faced by HCWs. Interestingly, some reacted to PMIEs in ways that protected them from moral dissonance. While not typical, some HCWs refused to abide by policies they morally disagreed with.

> *I mean we were happy to break those rules [visitation guidelines], we were on the verge of saying actually, we’re not going to let your father pass away on his own in a room without anybody around him..* (Peter, Nurse)

Notably, HCWs who exercised refusal tended to be older and either had worked for a longer time in their role, had returned from retirement during the pandemic, or held a position in middle management.

> *I’m getting old and cynical now so there’s definitely something about ageing that makes you a bit harder and you find it a bit easier to point things out both at work and in society in general.* (Harry, Nurse)

Similarly, when some HCWs were put in situations they did not feel trained to handle, they asserted the boundaries of their clinical competencies and committed to staying within them.

> *I would say I’m quite good at pushing back if someone asks me to work out of my competencies even my supervisor who I have had for five years she’s amazing, she’s a senior, she’s got qualifications and experience coming out everywhere she’s just an incredible level of knowledge, but I will flatly turn around to her and say I’m not doing that.* (Kathryn, Psychological therapist)

Where HCWs were pressured to do things that fell outside the remit of their role/training, they sometimes prevented moral dissonance by rationalising their actions in the context of the systemic constraints. This was only effective at preventing moral dissonance where no harm was caused to the patient.

> *So, there are times then when I thought I shouldn’t be doing this but there’s no one else to do it so at least someone is doing it. So at least I kind of had that and I knew that I wasn’t doing harm […]* (Harry, Nurse)

## 4. Psychological toll of PMIEs

### 4.i. Anger and guilt

This sub-theme encapsulates the emotional impact of moral distress.

Staff experienced anger towards the government for perceived mismanagement of COVID guidelines and for the under-funding of services, towards upper management in trusts related to feeling that the onus was wrongly put on staff to compensate for systemic failures by over-working themselves. This led to cynicism about organisational attempts to address staff well-being, which were seen not to address the root cause of the issue.

By contrast, guilt was experienced when HCWs felt personally responsible for letting others down. HCWs mainly felt this related to not being able to provide person-centred care during the pandemic, but interestingly HCWs voiced a degree of guilt by association, feeling complicit in a system they viewed as increasingly less equipped to service public need and provide high quality care.

> *I couldn’t do anything about it [patient becoming terminally ill due to delay in surgery] personally. So, in ways I felt guilt even though it wasn’t anything that I could do about it but still felt guilty as a member of the NHS for being almost a part in that.* (Chloe, Nurse)

### 4.ii. Disillusionment with the NHS

This sub-theme reflects a feeling of disillusionment with the perceived deterioration in standard of care provided by the NHS. While HCWs discontent surrounding patient backlogs, resource and staff shortages was present prior to the pandemic, many HCWs conveyed that their sense of disillusionment had grown with the exacerbation of these issues since the onset of COVID-19.

> *Nobody comes into nursing or mental health work or anything like that into that line of work to just be constantly frustrated. You know that there are shortages. It’s that helplessness, you can’t do anything about it.* (Owen, Nurse)

Many shared in the feeling that being prevented from providing holistic care undermined the degree of fulfilment they derived from their jobs: *“I’ve just lost that drive and ambition. It almost feels like at what point, there’s no end in sight*.” (Jamal, Doctor).

For some, this provided the impetus to leave their role and for others this spurred thinking about leaving the NHS entirely “*it’s a horrible feeling just not being able to do your job properly and it’s not particularly what I’m after so if there’s another place, I can be more useful then perhaps that’s what I’ll do.* (Brianna, Nurse).

### 4.iii. Deterioration of mental health

This sub-theme captures the adverse impact of moral distress on HCWs’ mental health. Deterioration in mental health was attributed to an accumulation of work-related moral distress, compounded by life stressors. Some HCWs described experiencing symptoms in the immediate aftermath of PMIE exposure, where others continued to grapple with long-term adverse mental health consequences of PMIEs.

> *I do genuinely feel injured […] the cumulative effect of that whole first wave of COVID and the redeployment and everything and not being listened to and feeling so deskilled just really hit me latently and that was very much PTSD like symptoms. So that affected me in terms of, well I mean I’ve been in recovery from alcohol problems since [year] and I had my first relapse.* (Conor, Healthcare Assistant)

This quote highlights the danger of the resurgence of old (or emergence of new) maladaptive coping strategies to combat moral distress. The most common impact of moral distress was heightened anxiety which led to sleep disturbances for several people.

> *I’ve had sleepless nights, some of those situations, some of the burden of carrying the staff and the negative emotions that they’ve had I’ve found extremely challenging and very upsetting* (Harry, nurse).

Anxiety appeared associated with several staff taking stress-related sick leave, and it manifested for one person in the form of panic attacks around the time of her night shifts, which resolved when she left her role. Presenteeism was described by Malaya who felt she could not take mental health related sick leave as she took an oath to protect her patients.

> *I’m emotionally, mentally, and spiritually drained. I’m tired every day. I didn’t want to feel like going to work but I did just because I took an oath to help people, right? Because I’m a nurse, I pushed myself. I encouraged myself to finish this redeployment.* (Malaya, Nurse)

Imelda related her anxiety to “*things being beyond what you are able to influence*” (Nurse). This sheds light on why some people’s anxiety improved when the threat of further PMIE exposure was removed (either due to changing role, end of redeployment, or reduced pressure on them at later stages of the pandemic), while those who continued to face an accumulation of PMIEs described persistent issues. For the latter HCWs, this was especially difficult to manage due to lack of opportunity to decompress and lack of access to supports.

> *I’m still waiting for occupational health at the hospital to get back to me after having self-referred three times.* (Jamal, Doctor)

## 5. Adaptively managing moral distress

This theme captures the two main strategies used by HCWs to adaptively manage their moral distress: confiding in a trusted other; and distracting from or switching off from thoughts of the event causing distress.

Many HCWs described the importance of being able to openly express their feelings of anger, guilt, and disillusionment, with a non-judgemental, empathic other. The source of this support varied for HCWs. Some confided in colleagues, whose similar experiences helped to normalise the moral distress they were feeling. Oliver (nurse) described using ‘*dark humour’* to discuss such experiences with colleagues to acknowledge and cope with the event. Mental health professionals spoke about utilising the supervisory relationship to help them externalise blame for adverse patient outcomes.

> *So, you are shifting the what we call the locus of control from internal focus, it’s not all my fault that I’ve not been able to support these people, to an external, well actually it’s the situation and maybe can. I guess it’s perspective in a word, it’s providing perspective.* (June, Psychological Therapist)

HCWs across settings mentioned the importance of being able to be heard in an ‘*unedited fashion’* and hearing the words ‘*it’s not your fault’,* highlighting the importance of feeling the acceptance and reassurance of another in the process of disclosure. For those without access to clinical supervision, reflective practice groups and trusted line managers sometimes fulfilled this role of providing assurance from a position of expertise or authority respectively. On the other hand, Bindu (pharmacist) caveated the utility of discussing experiences of moral distress, expressing the belief that it was unhelpful to continually revisit morally distressing memories through discussion, highlighting the balance to be struck between voicing feelings to promote dissolution rather than rumination.

Notably, some HCWs expressed that continued threat of PMIEs at work, and low perceived organisational support, precluded them from being able to process their experiences. Additionally, supervision was not necessarily seen to provide resolution, and for some acted merely a means to contain feelings.

> *[…] clinical supervision where you can offer that support and that restorative space is helpful. But it doesn’t necessarily address the ongoing difficulty, its kind of just supports you to deal with it.* (Nathan, Psychological Therapist)

Several HCWs described intentionally switching off from their emotions. HCWs found it helpful to engage in activities outside of work that demanded their attention in the moment (such as physical exercise, cooking, reading, meditating, spending time with family etc). These activities enabled temporary escape from difficult thoughts and helped to create separation between work and home life. However, it was not always possible for HCWs to create this mental separation, and this was especially difficult for many at night without the distraction of daily tasks, contributing to sleep disturbance.

### 5.i. Resolution of moral distress

A few staff members described being able to move past their experiences of moral distress both by shifting perspective of the situation as described above and regaining a sense of control by focusing on the good they could do professionally, moving forward. Notably, being able to move past such experiences was integrally linked to being able to learn from them to make changes to prevent repetition of the PMIE. For instance, following his experience witnessing family members of a dying patient being told to go home, Mike began carrying and displaying a lamination of the hospital visitation guidelines to colleagues on wards, to prevent patients’ relatives being told to leave the hospital, when permitted to visit. This enabled Mike to overcome his moral dissonance by focusing on the good he could do moving forward.

> *I’ve just resolved to keep my head down and get on with my bit of trying to do the little bit that I can. I’m not in charge of these people all I can do is try and influence them and role model and try and show what kindness looks like and what compassion is.* (Mike, Nurse)

Some experienced a more organic form of resolution that came with moving out of the environment they found morally injurious. For instance, HCWs whose redeployments ended meant they could return to comfortably working within their clinical competencies. Similarly, those who left their NHS Trust during the pandemic and moved to a more supportive working environment in a new Trust, were able to resolve feelings of betrayal they felt towards management in their previous Trust.

> *It did affect last year when I was still working there and then I had a, I was in a position where I was angry, I was frustrated with my employer etc. but I feel like now I don’t have that anymore.* (Amaya, Pharmacist)

Importantly, most HCWs continued to grapple with the adverse impact of their moral distress. There was a shared sense among participants that until organisations were perceived to acknowledge and meaningfully tackle the systemic issues giving rise to PMIE exposure, that experiences of moral distress would continue to accumulate, at a heavy personal cost.

> *The system demands that you make those kind of [ethically difficult] decisions. The more time you make those and the longer you are in that environment where you have to make those decisions, I guess the cumulative toll of that is significant.* (Matthew, Doctor)

## Discussion

This study sought to explore the experience and impact of PMIEs in a sample of frontline HCWs in England. Our findings proffer a cycle through which PMIEs accumulate, involving the perception of ongoing systemic NHS issues, ethical conflicts between staff and institutional priorities, and perceived deterioration of patient centred care. This study provides a novel contribution to the literature by demonstrating the interconnectedness of various PMIE sub-types among NHS HCWs and highlighting the impact of the pre-pandemic state of the NHS on staff’s ability to cope with PMIEs during the pandemic itself.

During crisis, the frequency of moral dilemmas encountered in healthcare may increase as the ethical climate shifts away from prioritising patient centred care to a more utilitarian, task-oriented model of care under increasing patient volume (Bayerle et al., 2022). This shift may indeed be a pragmatic response to deal with a public health crisis, despite severe disruption to normal working patterns (e.g. redeployment of staff and operating in novel environments and teams). However, our findings contribute to emerging evidence demonstrating the high personal cost of attempting to reconcile these incompatible value systems, resulting in burnout (Liberati et al., 2021) as well as feelings of helplessness, cynicism, disengagement from work, desire to change career direction(Patterson et al., 2021). We did not find that participants had been prepared for this shift in focus and despite purposive sampling of both participants who met clinical thresholds for adverse mental health outcomes as well as those for whom standardised reporting measures indicated no to mild symptoms; most participants discussed considerable adverse psychological impact related to moral distress. Notably, participant’s mental health status could not be meaningfully distinguished from their narrative accounts alone, highlighting the distress caused by PMIE exposure relates to both individuals who do and do not experience symptoms consistent with mental illness. Furthermore, this observation demonstrates the benefit of using qualitative methods to explore nuances in lived experience that may be missed using self-report instruments.

In the early stages of the pandemic, understanding of moral injury among HCWs focused on self-induced moral injury and the degree to which that might be expected to produce guilt and shame (Haller et al., 2020). However, evidence has begun to emerge suggesting experiences of betrayal (a form of other-induced MI) may be of particular concern among HCWs (Zerach & Levi-Belz, 2021), especially perceived betrayal by those in management positions in the NHS (French et al., 2021). Most recently, HCWs were shown to be nearly three times more likely to report ‘other-induced’ PMIEs compared to ‘self-induced’ (Nieuwsma et al., 2022). Consistently, we found experiences of betrayal were most often discussed among HCWs in our sample. Notably, our study demonstrates the dynamic relationship between perceived institutional betrayal and compromised patient care. We found HCWs were required to act in accordance with practices and policies they viewed as substandard. In turn, feeling forced to undermine their personal values contributed to feeling under-supported and betrayed. This is consistent with observations of combat veterans that in practice, other-oriented and self-oriented PMIEs are frequently intertwined (Shay, 2010).

While PMIE exposure often led to moral distress, this was not invariably the case. Evidence from the military suggests if moral stressors are not continuously experienced, then they may be attributed by individuals to situations or circumstances, and cognitively processed with no or only moderate psychosocial consequences (Litz et al., 2009). Indeed, several of our participants related their adverse mental health symptoms to the recurrence of PMIE exposure (as opposed to a once-off event), consistent with the argument that new situations may remind HCWs of their powerlessness in past situations, creating a crescendo effect potentially leading to moral injury or frank illness (Epstein & Delgado, 2010). In our sample, perceived organisational negligence surrounding cumulative PMIE exposure appeared key to engendering helplessness and frustration. Targets for intervention must therefore address perceptions that the NHS is unresponsive to staff and patient needs to reduce disillusionment with the system.

Considering the concept of moral repair, we found opportunities for disclosure and reflection with a trusted other facilitated adaptive management of moral distress. This is comparable to findings among combat veterans that disclosure was experienced as cathartic but insufficient to resolve moral distress; though some veterans reached resolution through formal psychological support to reframe the situation (Williamson et al., 2020). Important features of disclosure appear to be the opportunity to speak openly and honestly without judgement. Evidence suggests that efforts to promote a sense of righteousness and purpose may actually be counterproductive to the resolution of moral distress (MacLeish, 2018) and as such, future interventions should acknowledge the personal meaning individuals have ascribed to the event. Opportunities to instigate these conversations at a later date may be helpful, as strategies to distract from adverse emotions appear effective during crisis. Given that many HCWs felt resolution of moral distress was not possible without organisational change, a multi-facetted organisational strategy is warranted to both prevent excess PMIE exposure and to promote opportunities for resolution of moral distress. Accordingly, we propose a number of organisational recommendations to achieve this aim.

### Recommendations to prevent avoidable PMIE exposure

Firstly, NHS Trusts should proactively prepare staff to deal with PMIEs likely to be routinely encountered in their work. We suggest this is emphasised throughout professional training and refreshed periodically, especially following crisis events such as the pandemic. This training should be frank and focus on the likely impact of PMIEs and importantly ways of coping with them; it should have a positive focus highlighting that in most cases, any distress experienced will resolve if appropriately handled. Following on from the theme of ‘Strained working relationships with management’ where PMIEs were felt to accumulate due to issues with communication and conflicting priorities, we recommend NHS organisations ensure clear, transparent communication with staff of realistic expectations and targets, achievable in the context of staffing and resourcing constraints. To decrease the likelihood of staff ‘Feeling unable to fulfil adequate duty of care’ to patients, we recommend trusts consider distribute responsibility for clinical decision making, so that individual staff members do not bear the entire weight of responsibility for clinical outcomes. To address the perception of ‘Systemic NHS issues’ including failing to address staff concerns, we recommend trusts improve the responsiveness of channels designed to report workplace incidents (including witnessing violations in standards of care as well as to report instances of bullying or mismanagement). To more broadly combat staff feeling ‘ill equipped and under-supported’, we recommend trust leaders at all levels acknowledge the value discrepancy between person-centred care and the more task-oriented model of care necessitated in response to substantial increases in patient volume. Further, Trusts should seek to promote team discussion of how to address this conflict in public health crises. Finally, where PMIE exposure is unavoidable, we recommend trusts seek to promote adaptive management and resolution of moral distress by initiating reflective practice groups to help staff process their actions/inactions in the context of situational constraints and promote cognitive structuring to resolve potential feelings of guilt and self-blame.

### Strengths and limitations

This study has several important strengths. Firstly, this paper addresses an important knowledge gap surrounding the lived experience of HCWs with self-reported PMIE exposure in the NHS. Secondly, in keeping with qualitative epistemology, we recruited as inclusively as possible. To our knowledge, no other qualitative study on this subject (e.g. French et al., 2021) has achieved the range of demographic and occupational diversity of the current sample. Thus, an important strength of the study is the transferability of findings to HCWs who report PMIE-exposure.

Nevertheless, this study has its limitations. Inherent in all qualitative designs, inferences about causality between PMIE exposure and psychological distress are beyond the scope of the research. As this study was nested within a larger cohort study, participants were self-selected from an already self-selected sample and so were inclined to engage with research and willing to speak to people about their experience. Given that PMIE exposure was a criterion for inclusion in the current study, this study does not reflect the views and experiences of HCWs who were not exposed to any PMIEs during the pandemic. Thus, by virtue of the study design, our findings are not applicable to HCWs who did not experience PMIEs. For instance, findings from the NHS Staff survey indicate that 75.6% of staff respondents reported that care of patients/service users is their organisations top priority-in contrast to the experience of participants in the current study for whom this value discrepancy constituted a PMIE (NHS Staff Survey 2021 National Results Briefing, 2022). Furthermore, despite purposive recruitment of those exposed to MI by commission, this did not feature heavily in participants’ narratives. It is possible that this reflects a particular difficulty in discussing ones’ own transgressive acts relative to disclosure of one’s failure to act and experiences of betrayal. As moral injury is a broadly encompassing construct, future research may benefit from exclusive exploration of commission related PMIEs to enhance understanding of this dimension of moral injury. To limit under-reporting of such experiences, methods that allow anonymous responses (e.g. online qualitative survey designs) could be used, and recruitment materials should seek to normalise these experiences in times of crisis, and describe and explain the difficult feelings someone might expect in such circumstances.

### Implications

This study provides an in-depth insight into the experiences, views and needs of frontline HCWs routinely exposed to PMIEs during the COVID-19 pandemic. Studies to date have not included as diverse a sample, nor captured the current impact of PMIE accumulation across all waves of the pandemic. We found that the psychological toll of recurrent PMIEs is significant. This study extends the evidence base by demonstrating the interconnectedness of PMIE sub-types of betrayal and omission among NHS HCWs and highlights the urgent need to disrupt this cycle of exposure to better protect the mental wellbeing of staff and prevent organisational disengagement.

## Data Availability

 The data that support the findings of this study are available from the corresponding author (NG), upon reasonable request. The data have not been made publicly available due to the personal and sensitive content of participants experiences.

## Acknowledgements

We wish to acknowledge the National Institute of Health Research (NIHR) Applied Research Collaboration (ARC) National NHS and Social Care Workforce Group, with the following ARCs: East Midlands, East of England, South West Peninsula, South London, West, North West Coast, Yorkshire and Humber, and North East and North Cumbria. They enabled the set-up of the national network of participating hospital sites and aided the research team to recruit effectively during the COVID-19 pandemic. This paper is independent research supported by the National Institute for Health and Care Research ARC North Thames. The views expressed in this publication are those of the author(s) and not necessarily those of the National Institute for Health and Care Research or the Department of Health and Social Care.

The NHS CHECK consortium includes the following site leads: Sean Cross, Amy Dewar, Chris Dickens, Frances Farnworth, Adam Gordon, Charles Goss, Jessica Harvey, Nusrat Husain, Peter Jones, Damien Longson, Richard Morriss, Jesus Perez, Mark Pietroni, Ian Smith, Tayyeb Tahir, Peter Trigwell, Jeremy Turner, Julian Walker, Scott Weich, Ashley Wilkie.

The NHS CHECK consortium includes the following co-investigators and collaborators: Peter Aitken, Anthony David, Rosie Duncan, Cerisse Gunasinghe, Sam Gnanapragasam, Stephani Hatch, Daniel Leightley, Isabel McMullen, Paul Moran, Dominic Murphy, Martin Parsons, Catherine Polling, Alexandra Pollitt, Danai Serfioti, Chloe Simela, Charlotte Wilson Jones.

## Disclaimer

This funder had no role in study design, data collection, data analysis, data interpretation or writing of the report. This report is independent research supported by the National Institute for Health and Care Research ARC North Thames. The views expressed in this publication are those of the author(s) and not necessarily those of Manolo Blahnik International Limited, the NHS, the National Institute for Health and Care Research or the Department of Health and Social Care.

## Data availability statement

[The data that support the findings of this study are available from the corresponding author (NG), upon reasonable request. The data have not been made publicly available due to the personal and sensitive content of participants’ experiences.

## Appendices

## Appendix A

### Moral Injury Interview Schedule

<u>Group 1: Individuals who have reported symptoms consistent with PTSD, severe depression or</u> <u>severe anxiety (N=15)</u>

<u>Group 2: Individuals who have not reported symptoms consistent with PTSD, severe depression or</u> <u>severe anxiety (N=15)</u>

#### Introduction

This interview is about your exposure to potentially morally injurious events in the workplace while responding to the COVID-19 pandemic. Moral injury refers to the distress people experience, often in the form of shame, anger, guilt or disgust, when they see or carry out acts that clash with their deeply held moral or ethical beliefs and values. This may be a result of things you, or others, have done; things you or others did not do but you think should have happened; or from experiencing betrayal by those who you trusted or who should have been looking out for you.

Some examples of moral injury that have been recorded by NHS frontline staff to date include having to choose which of several sick COVID patients to ventilate due to resource constraints; not having the right medicines or equipment to save a person; putting non-COVID patients, or staff, at risk of contracting the virus due to inadequate PPE; feeling let down and unsupported by the NHS, government, or wider society. Of course, those are just examples, and your experience may be quite different. Does that all make sense?

Please do let me know if you have any questions at any point. The interview will last approximately 45 minutes and will ask you about your experience of events that potentially contribute to moral injury and the impact these events have had on you. I appreciate this may be a difficult topic to speak about. If at any point you’d like a break or to stop the interview we can pause and have a chat about how you’re feeling, reschedule to continue at another time, or stop completely. That being said, are you happy to begin the interview?

Could you please let me know your age

1. To begin, could you please tell me about your role(s) in the NHS during the pandemic?

- Has the nature of your job changed from what you were doing prior to the pandemic (e.g., redeployment).
- Who made the decision regarding the change in your role (was this decision made for you, did you decide/agree to it?)
- [If there was a change] How did you feel about the change in role?
2. Can you please tell me about how you felt working for the NHS before the pandemic? Has this has changed over the course of the pandemic and if so how and why?

- Have your relationships with colleagues/managers changed much since the pandemic?
- What about your relationship wi th people you are close to outside of the NHS like friends and family?
3. Thinking about your work during the pandemic, can you tell me about any experiences you had, witnessed, or learnt about, that clashed with your moral or ethical values(?).

- If they did, were these events a result of actions of you, or others? Please tell me a little about them?
- Did you experience a sense of betrayal from people you trusted, or who should have been looking out for you, during the pandemic, if so can you describe this briefly? (prompt for colleagues, management, organisation, healthcare staff you consulted, friends, family etc)
- On what occasions did you experience moral injury/events mentioned? (prompt: how frequent?)
- Roughly when did you experience the moral injury/injuries you described? (How was this similar to/different from your experience before the pandemic?)
4. What impact did this/these morally challenging events have on you?

- Emotional (anger, guilt, shame, disgust, sadness, etc)
- Psychological (i.e. have you felt sad, depressed, burned out, anxious because of it? Have you felt at all on edge since it happened; had any unpleasant dreams/disrupted sleep since then; have you
found yourself avoiding anything that reminds you of it?)
- Relational (impact on family life, social life, colleagues, team work)
- Occupational (impact on ability to work, intention to change role/Trust/leave healthcare altogether, sick leave, team morale, feeling supported at work)
- Spiritual (impact on their faith/world view or beliefs)
- Functional (ability to do daily tasks at work and personal life)
- Other
5. How long have these feelings/effects lasted?

- Do you still feel that way (constant vs come and go)
- How strong are the feelings you described?
6. Have the experiences you described influenced other areas of your life/life stressors (e.g. difficulties at home)?

- Has the pandemic created a backlog of work for you, and if so, has this impacted your ability to deal with the experiences you described?
- How have the morally challenging experiences you described changed your relationships to patients, if at all (more conflict, avoidance, less contact…)
7. What supports have you found helpful in coping with these morally challenging experiences? E.g.: -Informal (family friends, colleagues)

- Formal (psychological counselling, debriefing, time away from work etc)
- Personal (coping strategies, resilience, personality factors etc)
- Other
- Why have these things helped?
- How effective have these things been for you?
- Is there anything you think would be helpful to you right now?
8. What supports have you found unhelpful in coping with these morally challenging experiences?

- Informal (unsupportive family friends, colleagues)
- Formal (issues with psychological counselling, access to supports, organisational pressure)
- Personal (avoidant, anxious etc)
- Other
- Why have these things been unhelpful?
9. Has your trust done anything that makes dealing with exposure to potentially morally injurious events easier?

- Has your trust done anything that makes dealing with exposure to potentially morally injurious events more difficult?
10. Going forward, what measures could be taken by your trust to limit NHS staff experiences of potentially morally injurious events?

- any measures that you think might be helpful in future crises?
- any measures needed in general working life
- systemic changes within the NHS (such as additional training, added regulations, inputting reporting system in the aftermath of PMIES)?
11. Going forward, what do you think could be done differently to help NHS staff cope with the experience of moral injury, if it can’t be avoided?

- Why/how would these things help?
- Are you worried about anything that might be a morally challenging experience for you to deal with in future as we emerge from the pandemic? If so, can you tell me a bit about this?
- What impact might the changes you suggested have on your mental health, responding to future PMIES?

#### Interview Ending

Is there anything we haven’t yet covered that you feel is important to tell me? How did you find the interview?

Thank you for talking to me today. I really appreciate your time. The experiences you’ve shared today will be really helpful to understand the effects that moral injury can have on mental health. We hope this information will inform Trusts on how they can best support staff like you and your colleagues. If you would like, we can email you later in the year to update you on our progress and tell you about the key findings from our study.

We’ll also send your £25 gift voucher to the email address that we hold for you within 10 days, unless you’d like to provide me with a different email address that you’d prefer me to send it to. If you have any further comments or questions, you can email me at the email address.

